# The variability and performance of NHS England’s “Reason to Reside” criteria in predicting hospital discharge in acute hospitals in England. An observational study

**DOI:** 10.1101/2022.04.19.22274037

**Authors:** Elizabeth Sapey, Suzy Gallier, Felicity Evison, James Hodson, David McNulty, Katherine Reeves, Simon Ball

## Abstract

**Objectives:** NHS England (NHSE) advocates using “reason to reside” (R2R) criteria to generate a binary outcome, which supports discharge related clinical decision making. The proportion of patients without R2R and their rate of discharge are reported daily, by acute hospitals in England. R2R is however, not based upon an inter-operable standardised data model (SDM), nor has its performance been validated against its purpose. We aimed to understand the degree of inter- and intra-centre variation in R2R related metrics reported to NHSE, define a SDM implemented within a single centre Electronic Health Record to generate an eR2R, and evaluate its performance in predicting subsequent discharge.

**Design:** Retrospective observational cohort study using routinely collected health data.

**Setting:** 122 NHS Trusts in England for national reporting and an adult acute hospital in England for local reporting.

**Participants:** 6,602,706 patient-days were analysed using 3 months national data and 1,039,592 patient-days, using 3 years single centre data.

**Main outcome measures:** Variability in R2R related metrics reported to NHSE. Performance of eR2R in predicting discharge within 24 hours.

**Results:** There were high levels of intra and inter-centre variability in R2R related metrics (p<0.0001), but not in eR2R. Informedness of eR2R for discharge within 24 hours was low (J-statistic 0.09 – 0.12 across three consecutive years). In those remaining in hospital without eR2R, 61.2% met eR2R criteria on subsequent days (76% within 24 hours), most commonly due to increased NEWS2 (21.9%) or intravenous therapy administration (32.8%).

**Conclusions:** R2R related performance metrics are highly variable between and within acute Trusts in England. Although case-mix or community care provision may account for some variability, the absence of a SDM is a major barrier to meaningful interpretation of these metrics. The performance of eR2R based on two alternative SDM’s was poor, such that they could not meaningfully contribute to clinical decision making or evaluation of performance.

**Summary:** *What is known:* There is considerable pressure on hospital bed capacity and significant variation in hospital discharges with concerns raised about delays in discharge planning across National Health Service Trusts. To address this, the UK Government developed a policy and criteria to identify in-patients in whom discharge home, or to a less acute setting, should be considered. The criteria, called “reasons to reside” (R2R) have been promoted as a tool to improve discharge planning and are a mandated metric for central reporting. The performance of R2R has not been assessed.

*What this study adds:* This study suggests a low performance of the R2R criteria as a clinical tool to identify patients suitable for discharge, and questions its usefulness as a reported metric in its current form. There is significant intra and inter-centre variability in both the reported proportion of patients not meeting R2R criteria, and the proportion of patients not meeting R2R criteria who were later discharged. The proportion of patients not meeting R2R criteria correlates poorly with their rate of discharge over the subsequent 24 hours and the performance of the R2R criteria as dichotomous test to identify patients suitable for discharge is low. Further, the R2R criteria are not a stable phenomenon, with more than half of those who remain in hospital without R2R, subsequently acquiring a R2R during the admission.

## Introduction

In 2021 the UK Government published its policy and operating model for hospital discharge and community support within the National Health Service in England (NHSE) ^(1)^. This policy responded to concerns with respect to bed capacity during the COVID-19 pandemic and its sequelae, including the need to recover delayed activity and manage late presentation of other diseases.

The potential to release acute hospital beds was recognised in a report from the National Audit Office in 2016. This found that older patients no longer needing acute treatment, accounted for 2.7 million NHS hospital bed days per year^(2)^. There was evidence that lack of planning caused unnecessarily deferred discharge from hospital, which might contribute to inter-centre variation in length of stay. Collectively these data suggest that there are opportunities to improve the efficiency of healthcare delivery, especially since there is evidence that delayed discharge adversely affects individual patient experience and outcome^(3, 4)^.

The aforementioned policy included use of the previously developed discharge to assess model (referred to as “D2A”). This identifies a group of in-patients who no longer require acute admission, who nevertheless require additional care in their residence, whilst their baseline functional capacity returns or is established^(5)^. It also referred to use of a set of criteria to identify a group of in-patients in whom discharge home, or to a less acute setting, should be considered. These criteria have been referred to interchangeably, as “reasons to reside” (R2R), “right to remain” or “criteria to reside” (See Table 1a).

**Table 1.** Reason to Reside (R2R)

|  |  |
| --- | --- |
| 1 | Requiring ITU or HDU care |
| 2 | Requiring oxygen therapy / NIV |
| 3 | Requiring intravenous fluids |
| 4 | NEWS2 > 3 (clinical judgement required in persons with AF and/or chronic respiratory disease) |
| 5 | Diminished level of consciousness where recovery realistic |
| 6 | Acute functional impairment in excess of home/community care provision |
| 7 | Last hours of life |
| 8 | Requiring intravenous medication > bd (including analgesia) |
| 9 | Undergone lower limb surgery within 48 hours |
| 10 | Undergone thorax-abdominal/pelvic surgery within 72 hours |
| 11 | Within 24 hours of an invasive procedure? (with attendant risk of acute life-threatening deterioration) |

**Legend.** The policy and operating model for hospital discharge and community support within the National Health Service in England states that every person on every general ward should be reviewed on a twice daily ward round to determine whether they meet R2R. If the answer to each question is 'no', the policy states that active consideration for discharge to a less acute setting must be made (1).

Since April 2020, NHS hospitals have been required to provide daily reports on hospital discharges through the Strategic Data Collection Service (SDCS). These identify the numbers of people leaving hospital, to where, and the reasons for those remaining in hospital. The proportion of in-patients not meeting R2R criteria, and the proportion of patients without R2R discharged that day, are also reported. These metrics are considered to be measures of organisational and system efficiency.

R2R appears to have emerged heuristically from the clinical experience of those involved in its development. A series of questions are posed that might influence clinician behaviour and thereby prompt consideration of individual patients for discharge. However, there are no standardised data definitions, there has been no validation of the predictive power of R2R, no investigation of its role as clinical decision support at the patient level, or of its value in defining a subset of patients for the purposes of evaluating intra- or inter-centre performance. A further barrier to evaluating the performance of R2R is that there is no standard definition of patients who could be discharged from hospital against which to determine the predictive power of any set of criteria, were they to be defined. This lack of a reference standard limits, but does not preclude assessment of the validity of a clinical test, provided a ‘fair’ measure of performance can be defined^(6)^. The set of patients actually discharged in the subsequent 24 hours is one potentially ‘fair’ test of performance of R2R.

In the current study, we firstly show the degree of variation in R2R associated metrics reported across centres in England. Secondly, we propose precisely defined, inter-operable, data definitions corresponding to the elements of R2R. This allows for consistent, meaningful, generalisable analysis. Thirdly, we evaluate performance of R2R so defined in predicting discharge over the subsequent 24 hours.

## Methods

This study used unconsented, anonymous health data and all study activity was approved by the East Midlands–Derby REC (reference: 20/EM/0158) and was supported by PIONEER, the Health Data Hub in acute care. All studies activities followed the World Medical Association’s Declaration of Helsinki.

### National Data

National NHS England data was accessed via The UK Health Facts and Dimensions database^(7)^ for all reporting Trusts in England. Assessment of variability in national R2R reporting included data from 29^th^ November 2021 to 20^th^ February 2022. Table S1 of the online supplement provides the names of the Trusts whose data are presented anonymously. Data were collected daily during the censor period for 121 centres, yielding a total of 10,164 potential data points (centre-days). For each of these, the total number of occupied and unoccupied beds, and the number of patients with no right to reside were extracted. The numbers of patients with right to reside were then calculated by subtracting the number with no right to reside from the total number of occupied beds on that day. The number of General and Acute beds occupied in any given centre, on any given day (in-patients), was used as a surrogate for the number of patients eligible for evaluation using the R2R criteria. Review of the dataset found some missing, and potentially spurious data, which were excluded prior to analysis. This comprised instances where R2R data were not recorded (N=184 data points); where the total numbers of beds were either zero, missing, or clearly spurious (N=37 data points); or where there were more patients with no R2R than the total number of beds (N=3 data points).

### Local Data

In-depth analysis of R2R criteria were performed using data from the Queen Elizabeth Hospital Birmingham (QEHB). QEHB is a National Health Service (NHS), urban, adult, acute hospital in England which in 2019 had 1269 beds including 80 level 2/3 intensive care (ICU) beds, an Emergency Department that assesses >300 patients per day, and a mixed secondary and tertiary practice that includes all major adult specialities except for obstetrics and gynaecology. Bed numbers and in particular ICU bed numbers significantly increased in 2020 and 2021 at the peak of the COVID-19 pandemic. The electronic healthcare record (EHR) at QEHB (PICS, Birmingham Systems) contains time-stamped, structured records that include demography, location, admission and discharge, co-morbidities, physiological measurements supporting NEWS2 and Glasgow Coma Scale, operation noting, prescribing and investigations. The R2R criteria in Table 1 were mapped to computable definitions derived from the EHR (See Table 2), to generate an electronic R2R (eR2R). The OPCS Classification of Interventions and Procedures codes mapped to criterion 9-11 are described in Table S2 of the online supplement. The concept ‘acute functional impairment in excess of home/community care provision’, had no direct correlate. Safer Nursing Care Tool (SNCT) levels of care were however available^(8)^. SNCT level 2 and 3 correspond closely with the requirement for HDU or ICU^(9)^. Level 1a identifies patients requiring enhanced nursing reflecting acuity of illness and Level 1b identifies a group with increased nursing dependency. Level 1b is likely to include those who would and would not be considered to require ongoing care in acute hospital. SNCT level 1 was included in the definition of eR2R in two ways, including (eR2Rab) and excluding (eR2Ra) level 1b, to determine if this affected performance.

**Table 2.** Data definitions used to operationalise R2R for EHR.

|  | Flag if... | R2R criterion number |
| --- | --- | --- |
| On ITU HDU | listed as being in ITU or HDU ward | 1 |
| SNCT Level $\geq 2$ | Most recent SNCT level in previous 48 hours $\geq 2$ | 1 |
| SNCT Level 1a | Most recent SNCT level in previous 48 hours = 1a | 6 |
| SNCT Level 1b | Most recent SNCT level in previous 48 hours = 1b | 6 |
| Oxygen therapy/ NIV | oxygen administration or NIV<br>documented in observation chart within previous 24 hours | 2 |
| Intravenous fluids | iv fluid administration initiated in previous 24 hours or<br>variable rate insulin infusion administered in previous 24 hours | 3 |
| NEWS2 | if NEWS2 $> 3$ within last 24 hrs | 4 |
| Diminished consciousness | Glasgow Coma Scale value $\leq 12$ in last 24 hours | 5 |
| Last hours of life | comfort observation completed current<br>OR<br>End of Life medication bundle administered within last 24 hours | 7 |
| Intravenous prescription ≥ tds current (regular not prn) | IV medication prescribed within last 24 hours and frequency $\geq 3$ times per day for regular medication only | 8 |
| Intravenous medication administration ≥ tds within 24 hrs | IV medication administered $\geq 3$ times within last 24 hours | 8 |
| Lower limb surgery within 48hrs | Procedure with relevant OPCS codes in previous 48 hours | 9 |
| Thorax-abdominal-pelvic surgery with 72hrs | Procedure with OPCS relevant codes in previous 72 hours | 10 |
| Invasive procedure within 24hrs | Procedure with OPCS relevant codes in previous 24 hours | 11 |

The primary analysis of eR2R was for patients who had been in hospital for more than twenty-four hours at midnight. Discharge over the course of the subsequent twenty-four hours was evaluated. Secondary analyses were undertaken for the set of patients in a bed at 08:00 and at 16.00 to define any change in eR2R performance in these different cross sections of the in-patient population. Three calendar years were analysed separately, to assess the effects of the COVID19 pandemic.

### Statistics

Initially, daily numbers of patients with right to reside, quantified both as absolute numbers and a proportion of the total number of beds, were plotted for national centres and used to calculate between-centre and within-centre variation. These data are analysed as beds occupied at the specified time of day, where the bed inherits the demographics, comorbidities, and other qualities of the occupying patient. This then represents the in-patient population in cross-section. This approach was replicated in the local analysis of eR2R: the term patient-day was used to refer to a bed with the qualities of the occupying patient at the time of the analysis. The in-patient population is described as means of patient-days thereby representing a cross-section of the group. The performance of eR2R as a predictor of remaining in hospital (or absence of eR2R as a predictor of discharge) was reported as a True Positive Rate (TPR) and True Negative Rate (TNR), Positive Predictive Value (PPV), Negative Predictive Value (NPV) and Youden’s J statistic (TPR+TNR-1), where positive is remains in hospital and negative is discharge from hospital within 24 hours.

Normally distributed variables are reported as arithmetic means ± standard deviations, with medians and ranges used otherwise. Between-centre variation was assessed by ANOVA. This included a model accounting for day of the week as a fixed effect and the centre as a random effect. All analyses were performed using IBM SPSS 22 (IBM Corp. Armonk, NY), with p<0.05 deemed to be indicative of statistical significance throughout

### Patient and public involvement

Patients and public members have been involved in this study from inception. The research question and topic were agreed following patient/public discussion groups about NHSE discharge policies. Patients/public reviewed the data fields included in the study for both national and local data, with the PIONEER Data Trust Committee providing support for the project (a group of patient/public members who review studies using health data). A patients/public group have reviewed the results and have written a lay summary for study dissemination to patient groups. These will be translated into locally prevalent languages in Birmingham.

## Results

### R2R reporting in England Nov 20-Feb 21

Across 10,164 available centre-days, accounting for 6,602,706 patient-days, the number of patients reported without R2R as a proportion of in-patients, varied significantly between centres (p<0.0001). Individual centre means ranged from 6.7% ± 2.5% to 59.9% ± 13.8% (Figure 1a). There was also marked within-centre variation (Figure 1a), with coefficients of variation (CV) ranging from 8.2% up to 59.3%. Of patients not meeting R2R criteria, the proportion discharged over the following 24 hours, varied significantly between centres (p<0.0001). Individual centre means ranged from 14.0% ± 7.4% to 85.8% ± 25.2% (Figure 1b). There was also marked within centre variation, with coefficients of variation ranging from 6.4% up to 83.2%. This variation was not significantly altered by accounting for effects of day of the week (Fig S1). The proportion of patients without R2R and the proportion of that group discharged within 24 hours, were only weakly correlated (R^2^=0.12; Figure S2 of the online supplement).

**Figure 1.**
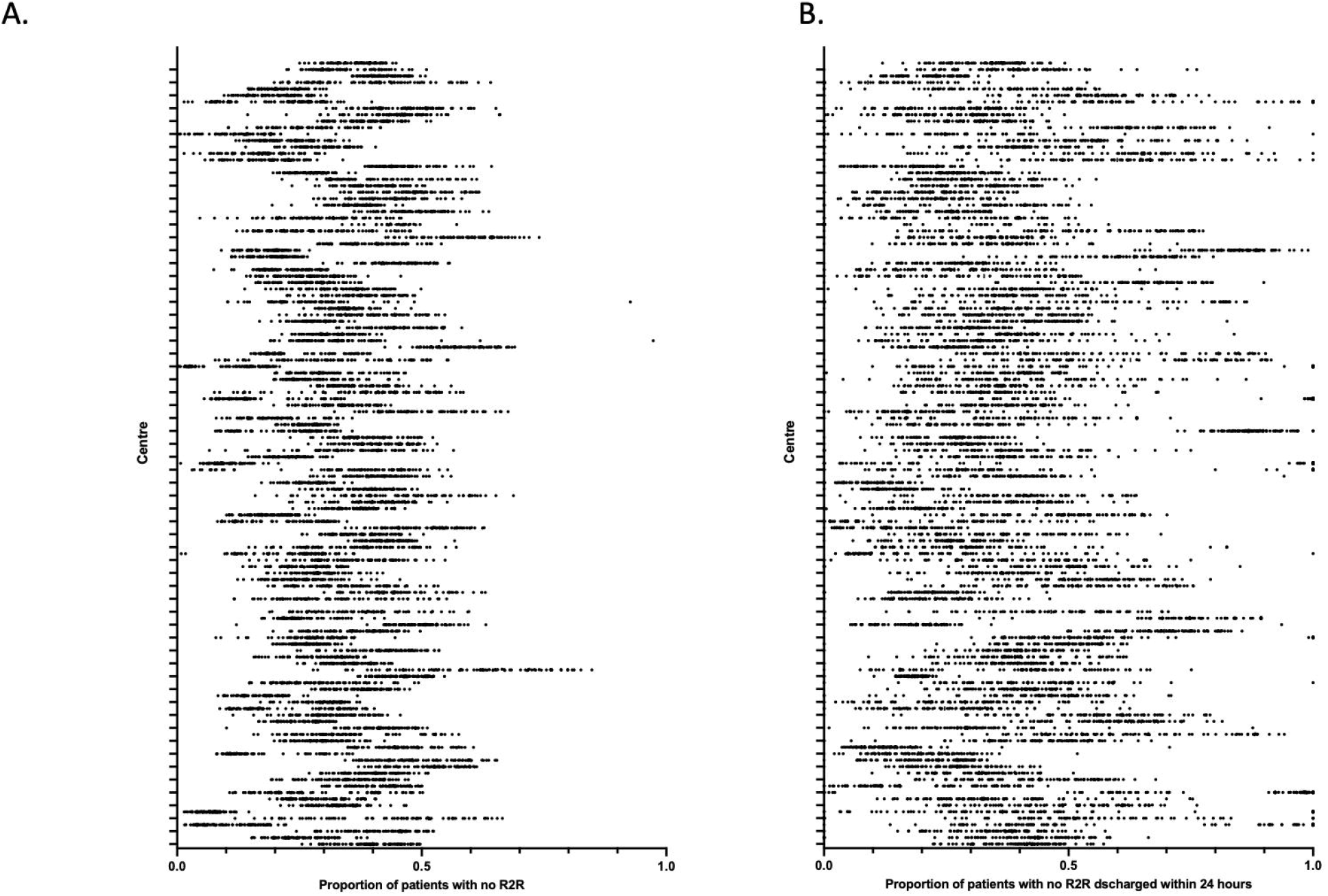
National reporting of R2R criteria. The proportion of patients with no R2R (Figure 1A) and of that group the proportion of patients discharged within 24 hours (Figure 1B) reported to SDCS from 29 Nov 2021 – 20 Feb 2022 across 121 centres. Each dot represents result for a single centre-day.

### Performance of eR2R at QEHB

#### Patients and admissions

Standardised definitions corresponding to the elements of R2R (Table 2) were used to analyse data from QEHB, on 1,214,480 in-patient days, between 01 Jan 2019 – 31 Dec 2021. The demographic and clinical details of that population are summarised in Table 3 which also shows that those meeting the definition of eR2Rab were older and more likely to have one or more co-morbidities than those who did not. Variation in the daily number of patients with or without an eR2R is shown in Figure S3 of the online supplement.

**Table 3.**
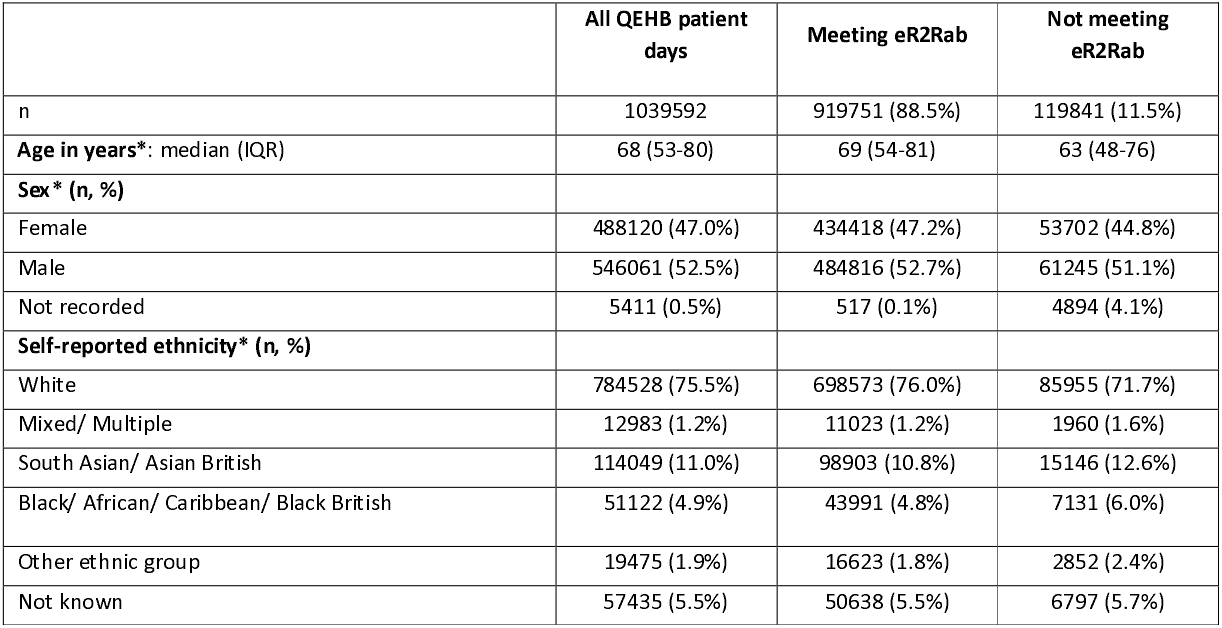
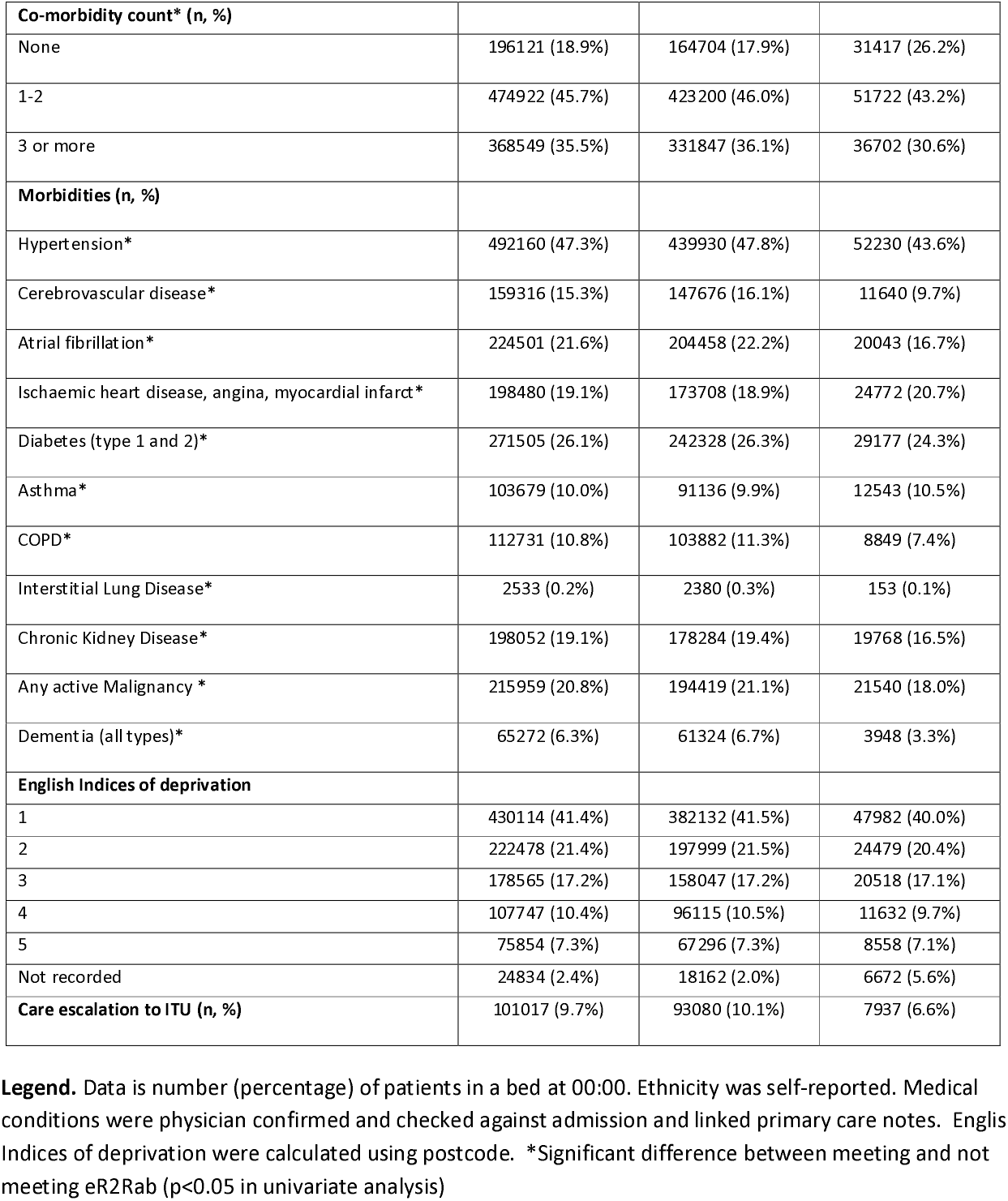
Demographics of patients meeting and not meeting R2R criteria on presentation to QEHB in the censor period.

### Criteria contributing to eR2R

Given the potential for the COVID19 pandemic to affect the performance of R2R, calendar years were analysed separately. The number of patients meeting any given eR2R criterion are shown in Table 4a. The progressive contribution of different elements of the definition of eR2R assessed daily in a modified Consort table, are summarised in Table 4b. The proportion of patients not meeting eR2R criteria exhibited relatively little day to day variation in 2019 (eR2Rab, CV = 11.2%; eR2Ra, CV = 6.3%), although somewhat higher in the context of case mix variation consequent upon peaks of patients admitted with COVID-19 in 2020 (eR2Rab, CV = 23.3%; eR2Ra, CV = 14.4%) and 2021 (eR2Rab, CV=17.1%; eR2Ra, CV = 9.9%). The criteria contributing most to eR2R status included acuity level (NEWS2 >3), SNCT level nursing requirement, being on an intensive care unit and requiring intravenous medications or fluids.

**Table 4a.** The number (percentage) of patient-days on which each eR2R data definition was met.

| Year | 2019 | 2020 | 2021 |
| --- | --- | --- | --- |
| Criterion | n (%) | n (%) | n (%) |
| ICU | 22899 (6.1) | 20326 (6.7) | 21305 |
| TAP surgery 72Hrs | 3783 (1.0) | 3010 (1.0) | 3974 |
| Lower limb surgery 48Hrs | 285 (0.1) | 252 (0.1) | 221 (0.1) |
| Invasive surgery 24Hrs | 1861 (0.5) | 1613 (0.5) | 1988 (0.6) |
| NEWS2 > 3 24hrs | 93501 (24.8) | 85123 (27.9) | 97722 (27.3) |
| O2 Treatment 24Hrs | 77949 (20.7) | 69355 (22.7) | 77202 (21.6) |
| Insulin Infusion 24Hrs | 10951 (2.9) | 10860 (3.6) | 12496 (3.5) |
| IV Fluids 24Hrs | 79802 (21.2) | 71376 (23.4) | 80246 (22.4) |
| IV medication administered in last 24hrs >= tds | 95034 (25.2) | 81174 (26.6) | 91573 (25.6) |
| IV medication prescribed in last 24Hrs >= tds | 21543 (5.7) | 17866 (5.9) | 19249 (5.4) |
| SNCT Dependency 1a, 2, 3 | 99139 (26.3) | 72226 (23.7) | 88832 (54.8) |
| COMA Score <=12 in last 24Hrs | 6594 (1.8) | 6448 (2.1) | 6664 (1.9) |
| End of Life care definition met in last 24Hrs | 5359 (1.4) | 4747 (1.6) | 5075 (1.4) |
| SNCT Dependency 1b | 172659 (45.8) | 160380 (52.5) | 179527 (50.2) |
| Total number of patient days | 376684 | 305254 | 357654 |

**Legend.** The number (percentage) of patient days on which each eR2R definition was met. The population was in-patients at 24.00 with length of stay ≥ 24 hours.

**Table 4b.** Daily mean contribution of each eR2R data definition in a phased analysis (modified Consort diagram)

| Year | 2019 | 2020 | 2021 |
| --- | --- | --- | --- |
| Criterion | Mean % (SD) | Mean % (SD) | Mean % (SD) |
| TAP surgery 72Hrs | 0.7% (0.35) | 0.7% (0.37) | 0.8% (0.45) |
| Lower limb surgery 48Hrs | 0.1% (0.07) | 0.1% (0.11) | 0.1% (0.08) |
| Invasive surgery 24Hrs | 0.2% (0.15) | 0.2% (0.18) | 0.2% (0.15) |
| NEWS2 > 3 24hrs | 24.2% (2.28) | 27.5% (3.82) | 26.6% (3.64) |
| O2 Treatment 24Hrs | 4.0% (0.61) | 3.9% (0.72) | 3.6% (0.68) |
| Insulin Infusion 24Hrs | 0.5% (0.24) | 0.6% (0.28) | 0.5% (0.23) |
| IV Fluids 24Hrs | 8.8% (1.09) | 9.5% (1.37) | 9.6% (1.24) |
| IV Medication Admin 24Hrs ≥ tds | 7.7% (1.05) | 7.4% (1.29) | 7.5% (1.17) |
| IV Medication Prescribed 24Hrs | 0.7% (0.28) | 0.6% (0.29) | 0.6% (0.27) |
| Dependency 1a, 2, 3 | 8.8% (1.42) | 6.7% (1.21) | 7.8% (1.12) |
| COMA Score ≤12 24Hrs | 0.0% (0.05) | 0.0% (0.08) | 0.0% (0.06) |
| End of Life 24Hrs | 0.5% (0.24) | 0.4% (0.27) | 0.4% (0.19) |
| Dependency 1b | 24.5% (1.88) | 25.5% (3.53) | 25.3% (2.59) |

| Criterion | Mean % (SD) | Mean % (SD) | Mean % (SD) |
| --- | --- | --- | --- |
| ICU | 6.1% (0.44) | 7.1% (3.10) | 6.0% (2.16) |
| TAP surgery 72Hrs | 0.7% (0.35) | 0.7% (0.37) | 0.8% (0.45) |
| Lower limb surgery 48Hrs | 0.1% (0.07) | 0.1% (0.11) | 0.1% (0.08) |

### Informedness of eR2R for discharge in the next 24 hours

For the outcome discharge (remain -) / no discharge (remain +) within 24 hours, across the 3 different years, the eR2Ra TPR lay between 0.63 and 0.65, TNR between 0.46 and 0.47, the PPV was 0.91 and NPV between 0.12 and 0.15; the eR2Rab TPR lay between 0.88 and 0.91, TNR between 0.18 and 0.24, the PPV between 0.90 and 0.91 and NPV between 0.18 and 0.20 (Table 5). The J statistic for both definitions lay between 0.09-0.12. In secondary analyses based upon the in-patient population at 08.00 and at 16.00 the J-statistic ranged between 0.10-0.14 and 0.10-0.15 respectively (Tables S3a and S3b of the online supplement).

**Table 5.**
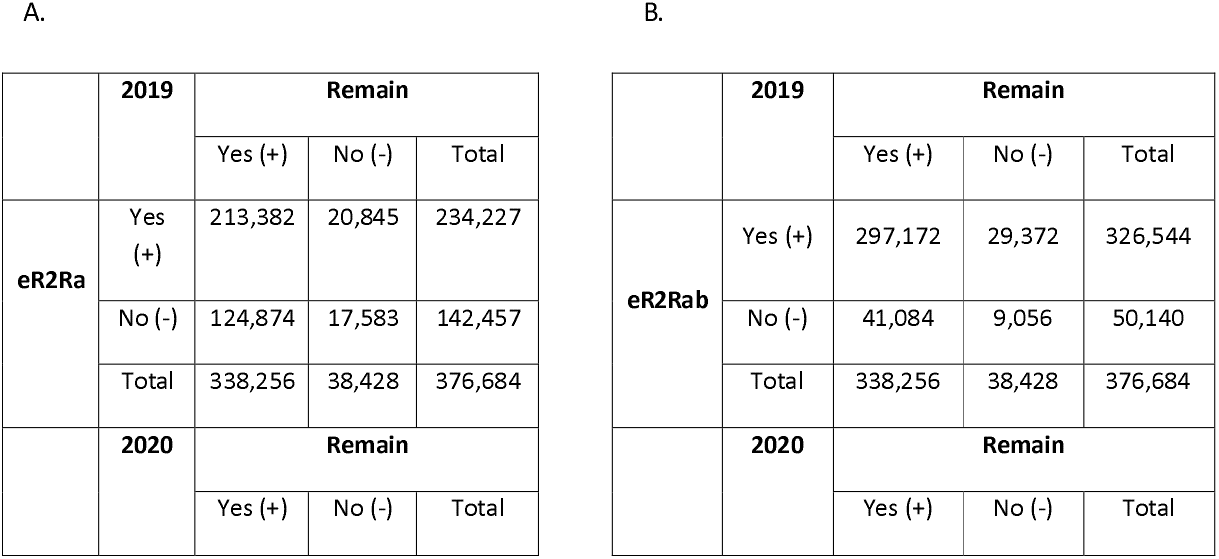
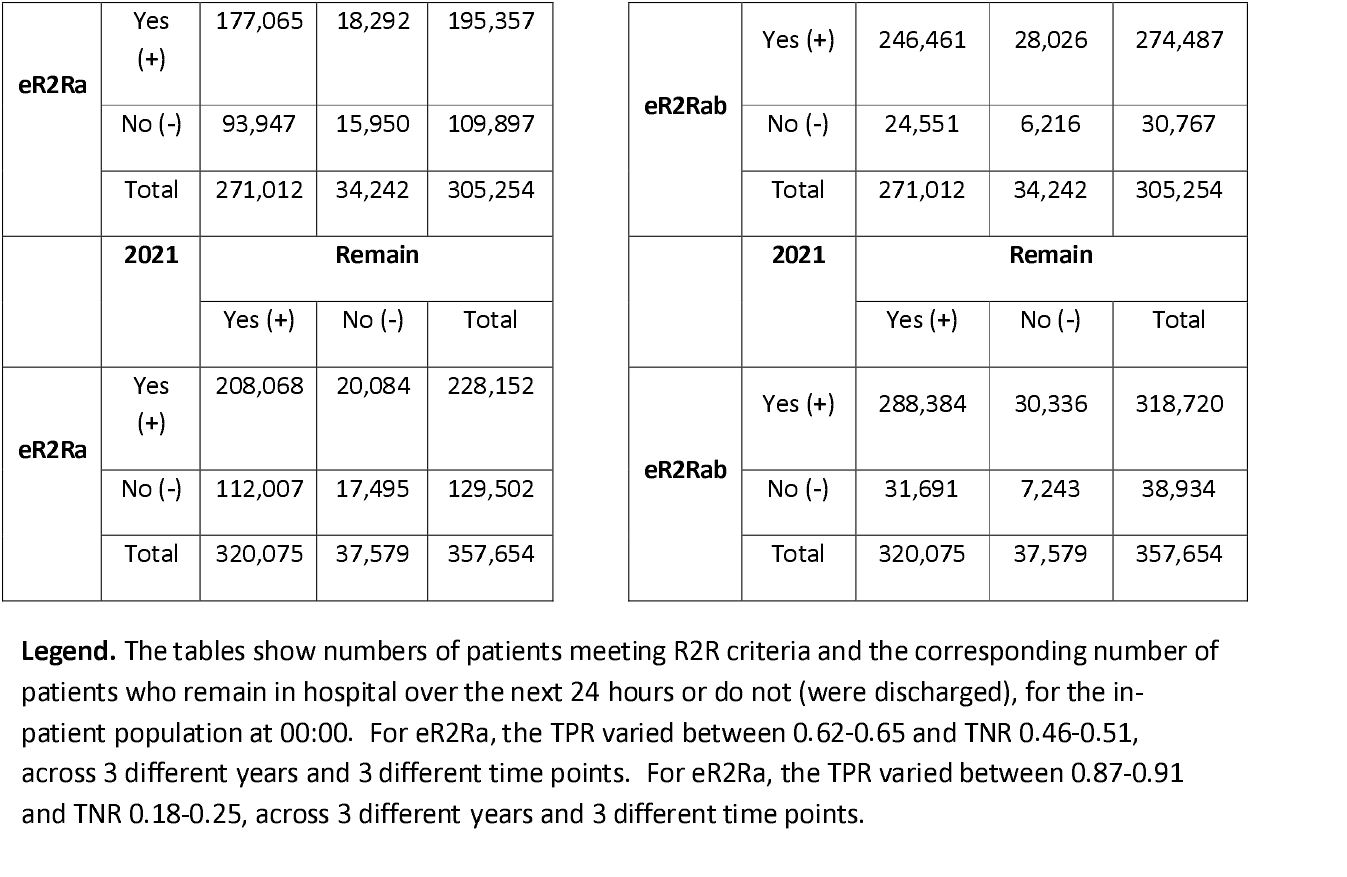
Contingency tables showing the number of patients meeting criteria for (A) eR2Ra and (B) eR2ab.

### In-patients not meeting eR2R

The demographic and clinical details of patient who did not meet the eR2Rab definition, stratified by discharge in the subsequent 24 hours are shown in Table S4 of the online supplement. For patient-days on which discharge occurred within 24 hours, there was significantly higher representation of those with no documented co-morbidities 29.2% vs 24.0% (p<0.0001). In those that remained in hospital, 61.2% met eR2R criteria on subsequent days (76% within the next 24 hours). Of all those that remained, 21.9% acquired a NEWS2 > 3, 32.8% received iv fluids or drugs > 3 times / day and 1.9% were admitted to ICU.

## Discussion

Assessment of an individual patient’s R2R has been promoted as a tool to improve the identification of those who could be discharged from acute hospitals in England. The proportion of in-patients with R2R and their rate of discharge has then been used to evaluate the operational efficiency of acute hospitals and their adjacent health and social care system^(1, 5)^. This paper presents findings to suggest that as currently constituted, R2R is in fact of limited value for these purposes.

The high levels of variation in R2R related metrics, within and between centres in England, has been attributed to variation in case mix and operational efficiency^(10)^. However, such extremes of variation are not observed in other metrics that use established data standards. Furthermore, the proportion of patients not meeting R2R criteria correlates poorly with their rate of discharge over the subsequent 24 hours, whereas one might anticipate that such closely related measures of operational efficiency would reflect one another. These findings are most obviously accounted for by the fact that R2R does not constitute a semantic data model. It is therefore susceptible to differing interpretation by individuals and centres. This applies to all the concepts described by R2R, but most obviously those that are necessarily subjective, such as ‘acute functional impairment in excess of home/community care provision’ and ‘diminished level of consciousness where recovery is realistic’^(11, 12)^.

We therefore developed machine readable data definitions corresponding to each concept, allowing consistent analysis of R2R at scale, using data derived from the EHR in our centre. The SNCT is a global nursing assessment of acuity and dependency that was developed to guide workforce deployment. It is regularly recorded within the EHR at our centre. Because Level 1b describes a group of patients who are highly dependent upon nursing care for daily activities, this was mapped onto the R2R concept ‘acute functional impairment in excess of home/community care provision’. However, since the definition of level 1b could include a group of patients suitable for discharge to a less acute setting, two definitions or eR2R were tested, with and without SNCT 1b. Our analysis is therefore likely to represent two extremes of inclusion of patients with acute functional impairment.

Within centre variation in eR2R was low, consistent with it minimising individual interpretation of each data element. eR2R was a poor predictor of discharge within 24 hours^(13)^. Youden’s Index was consistently <0.15 across 3 calendar years, 3 different times of day and two eR2R definitions. For a dichotomous test such as eR2R, a Youden’s Index >0.50 is generally considered the empirical benchmark for a test to support clinical decision making^(14)^. eR2R is therefore unsuited to the provision of clinical decision support tool for discharge. It does not define a sub-population on which to assess discharge performance^(15)^. The limitations of R2R are not entirely surprising, given the need to interpret concepts that are not semantically defined. Although addressed by eR2R, it nevertheless remains a simple series of binary responses to questions that have not been validated for the purpose of discharge prediction. For example, NEWS2 was validated as an acuity score to quantify physiological instability on initial presentation to hospital^(16)^. It was not developed and has not been validated, as a triage tool to assess fitness to leave hospital, at any threshold.

Importantly, more than half of those who remain in hospital without eR2R, subsequently acquired eR2R. This group of patients were older and had multiple long-term health conditions, suggesting that there were clinical grounds for that decision, albeit undefined. This sub-population requires further study.

There are limitations to our analysis. The eR2R was assessed in only one centre, albeit one that serves a diverse, multi-ethnic, urban population, in which more than 1.2 million patient days were assessed. Patients admitted for < 24 hours at the time of analysis were excluded, to allow clinical decisions to be made and executed. The first day post-admission is a highly dynamic situation, with frequent clinical review; a setting in which this embodiment of clinical decision support is arguably less relevant. Another, more intrinsic problem, is that there is no gold standard by which to define all patients suitable for discharge, so that actual discharge was used as a fair test when evaluating the performance of eR2R^(17)^. This assumes that patients actually discharged are part of a continuous population of all those who could be discharged. It is also the case that each eR2R data element could be defined in different ways, however each definition would relate to that used, so that the performance of one model would be informed by the other. For example, the 24-hour retrospective time horizon for most evaluations could be altered, but the later model would relate directly to the former.

Development of artificial intelligence-based systems to inform clinical decision making, has highlighted the importance of test validation and evaluation, within its intended setting. Stringent evaluation is equally important for clinical decision support tools, even when the underlying logic can be explained. The effects of embedding new care pathways or tools within clinical service delivery, without appropriate evaluation, are increasingly described. There is significant opportunity for unintended consequences to arise from the implementation of poorly considered clinical decision support^(18)^, particularly when there is competition for clinical resource. This has been recently discussed for NEWS2^(19)^, sepsis alerting and COVID-19 virtual wards^(20)^. R2R has been endorsed and adopted but without validation or consideration of the unintended consequences of its application. This is not to contend that a significant number of in-patients could not be discharged earlier, simply that there is no evidence that R2R can support clinical decision making. The collective limitations of R2R identified are likely to account for variation in nationally reported metrics which are difficult to explain.

Our study highlights the need for reproducible standardised data definitions to support both implementation and validation of any tool that purports to support clinical decision making. Further research should focus on building, validating and refining tools to inform clinical decisions.

## Data Availability

The anonymised dataset used for analysis is available upon reasonable request from the PIONEER Data Hub on submission of a data request form, see www.pioneerdatahub.co.uk for a copy of the form and processes for data access.

## Acknowledgements and funding

This work was funded by HDR-UK and was supported by the PIONEER Data Hub (see www.pioneerdatahub.co.uk). It was supported by the PIONEER patient and public advisory group and Data Trust Committee.

## Transparency statement

Professor Ball (the manuscript’s guarantor) affirms that the manuscript is an honest, accurate, and transparent account of the study being reported; that no important aspects of the study have been omitted; and that any discrepancies from the study as originally planned (and, if relevant, registered) have been explained.

## Author contribution

Simon Ball and Elizabeth Sapey conceived the study, Felicity Evison, James Hodson, David McNulty, Katherine Reeves and Suzy Gallier conducted data analysis. Elizabeth Sapey wrote the first draft of the study. All authors contributed to the study manuscript. Simon Ball is senior author and manuscript guarantor.

