## Supplementary material for "The variability and performance of NHS England’s “Reason to Reside” criteria in predicting hospital discharge in acute hospitals in England. An observational study": Online supplement figures and tables

**Online Tables**

| National Trusts | |
|---|---|
| •Airedale NHS Foundation Trust<br>•Ashford And St Peter's Hospitals NHS Foundation Trust<br>•Barking, Havering And Redbridge University Hospitals NHS Trust<br>•Barnsley Hospital NHS Foundation Trust<br>•Barts Health NHS Trust<br>•Bedfordshire Hospitals NHS Foundation Trust<br>•Blackpool Teaching Hospitals NHS Foundation Trust<br>•Bolton NHS Foundation Trust<br>•Bradford Teaching Hospitals NHS Foundation Trust<br>•Buckinghamshire Healthcare NHS Trust<br>•Calderdale And Huddersfield NHS Foundation Trust<br>•Cambridge University Hospitals NHS Foundation Trust<br>•Chelsea And Westminster Hospital NHS Foundation Trust<br>•Chesterfield Royal Hospital NHS Foundation Trust<br>•Countess Of Chester Hospital NHS Foundation Trust<br>•County Durham And Darlington NHS Foundation Trust<br>•Croydon Health Services NHS Trust<br>•Dartford And Gravesham NHS Trust<br>•Doncaster And Bassetlaw Teaching Hospitals NHS Foundation Trust<br>•Dorset County Hospital NHS Foundation Trust<br>•East And North Hertfordshire NHS Trust<br>•East Cheshire NHS Trust<br>•East Kent Hospitals University NHS Foundation Trust<br>•East Lancashire Hospitals NHS Trust<br>•East Suffolk And North Essex NHS Foundation Trust<br>•East Sussex Healthcare NHS Trust<br>•Epsom And St Helier University Hospitals NHS Trust<br>•Frimley Health NHS Foundation Trust<br>•Gateshead Health NHS Foundation Trust<br>•George Eliot Hospital NHS Trust<br>•Gloucestershire Hospitals NHS Foundation Trust<br>•Great Western Hospitals NHS Foundation Trust<br>•Guy's And St Thomas' NHS Foundation Trust<br>•Hampshire Hospitals NHS Foundation Trust<br>•Harrogate And District NHS Foundation Trust<br>•Homerton University Hospital NHS Foundation Trust<br>•Hull University Teaching Hospitals NHS Trust<br>•Imperial College Healthcare NHS Trust<br>•Isle Of Wight NHS Trust<br>•James Paget University Hospitals NHS Foundation Trust<br>•Kettering General Hospital NHS Foundation Trust<br>•King's College Hospital NHS Foundation Trust<br>•Kingston Hospital NHS Foundation Trust<br>•Lancashire Teaching Hospitals NHS Foundation Trust | •Salisbury NHS Foundation Trust<br>•Sandwell And West Birmingham Hospitals NHS Trust<br>•Sheffield Teaching Hospitals NHS Foundation Trust<br>•Sherwood Forest Hospitals NHS Foundation Trust<br>•Somerset NHS Foundation Trust<br>•South Tees Hospitals NHS Foundation Trust<br>•South Tyneside And Sunderland NHS Foundation Trust<br>•South Warwickshire NHS Foundation Trust<br>•Southport And Ormskirk Hospital NHS Trust<br>•St George's University Hospitals NHS Foundation Trust<br>•St Helens And Knowsley Teaching Hospitals NHS Trust<br>•Stockport NHS Foundation Trust<br>•Surrey And Sussex Healthcare NHS Trust<br>•Tameside And Glossop Integrated Care NHS Foundation Trust<br>•The Dudley Group NHS Foundation Trust<br>•The Hillingdon Hospitals NHS Foundation Trust<br>•The Newcastle Upon Tyne Hospitals NHS Foundation Trust<br>•South Warwickshire NHS Foundation Trust<br>•Southport And Ormskirk Hospital NHS Trust<br>•St George's University Hospitals NHS Foundation Trust<br>•St Helens And Knowsley Teaching Hospitals NHS Trust<br>•Stockport NHS Foundation Trust<br>•Surrey And Sussex Healthcare NHS Trust<br>•Tameside And Glossop Integrated Care NHS Foundation Trust<br>•The Dudley Group NHS Foundation Trust<br>•The Hillingdon Hospitals NHS Foundation Trust<br>•The Newcastle Upon Tyne Hospitals NHS Foundation Trust<br>•The Princess Alexandra Hospital NHS Trust<br>•The Queen Elizabeth Hospital, King's Lynn, NHS Foundation Trust<br>•The Rotherham NHS Foundation Trust<br>•The Royal Wolverhampton NHS Trust<br>•The Shrewsbury And Telford Hospital NHS Trust<br>•Torbay And South Devon NHS Foundation Trust<br>•United Lincolnshire Hospitals NHS Trust<br>•University College London Hospitals NHS Foundation Trust |

| | |
|---|---|
| •Leeds Teaching Hospitals NHS Trust<br>•Lewisham And Greenwich NHS Trust<br>•Liverpool University Hospitals NHS Foundation Trust<br>•London North West University Healthcare NHS Trust<br>•Maidstone And Tunbridge Wells NHS Trust<br>•Manchester University NHS Foundation Trust<br>•Medway NHS Foundation Trust<br>•Mid And South Essex NHS Foundation Trust<br>•Mid Cheshire Hospitals NHS Foundation Trust<br>•Mid Yorkshire Hospitals NHS Trust<br>•Milton Keynes University Hospital NHS Foundation Trust<br>•Norfolk And Norwich University Hospitals NHS Foundation Trust<br>•North Bristol NHS Trust<br>•North Cumbria Integrated Care NHS Foundation Trust<br>•North Middlesex University Hospital NHS Trust<br>•North Tees And Hartlepool NHS Foundation Trust<br>•North West Anglia NHS Foundation Trust<br>•Northampton General Hospital NHS Trust<br>•Northern Care Alliance NHS Foundation Trust<br>•Northern Devon Healthcare NHS Trust<br>•Northern Lincolnshire And Goole NHS Foundation Trust<br>•Northumbria Healthcare NHS Foundation Trust<br>•Nottingham University Hospitals NHS Trust<br>•Oxford University Hospitals NHS Foundation Trust<br>•Portsmouth Hospitals University National Health Service Trust<br>•Royal Berkshire NHS Foundation Trust<br>•Royal Cornwall Hospitals NHS Trust<br>•Royal Devon And Exeter NHS Foundation Trust<br>•Royal Free London NHS Foundation Trust<br>•Royal Surrey County Hospital NHS Foundation Trust<br>•Royal United Hospitals Bath NHS Foundation Trust | •University Hospital Southampton NHS Foundation Trust<br>•University Hospitals Birmingham NHS Foundation Trust<br>•University Hospitals Bristol And Weston NHS Foundation Trust<br>•University Hospitals Coventry And Warwickshire NHS Trust<br>•University Hospitals Dorset NHS Foundation Trust<br>•University Hospitals Of Derby And Burton NHS Foundation Trust<br>•University Hospitals Of Leicester NHS Trust<br>•University Hospitals Of Morecambe Bay NHS Foundation Trust<br>•University Hospitals Of North Midlands NHS Trust<br>•University Hospitals Plymouth NHS Trust<br>•University Hospitals Sussex NHS Foundation Trust<br>•Walsall Healthcare NHS Trust<br>•Warrington And Halton Teaching Hospitals NHS Foundation Trust<br>•West Hertfordshire Hospitals NHS Trust<br>•West Suffolk NHS Foundation Trust<br>•Whittington Health NHS Trust<br>•Wirral University Teaching Hospital NHS Foundation Trust<br>•Worcestershire Acute Hospitals NHS Trust<br>•Wrightington, Wigan And Leigh NHS Foundation Trust<br>•Wye Valley NHS Trust<br>•Yeovil District Hospital NHS Foundation Trust<br>•York And Scarborough Teaching Hospitals NHS Foundation Trust |

**Table S1. The names of the Hospital Trusts included in the national R2R reporting analysis**
**Legend.** Data is presented anonymously

| Procedure | OPCS Codes |
|---|---|
| Lower limb surgery within 48hrs | See Online supplement xls. |
| Thorax-abdominal-pelvic surgery with 72hrs | See Online supplement xls. |
| Invasive procedure within 24hrs | See Online supplement xls. |

**Table S2. Codes used to identify surgical interventions.**
**Legend**. OPCS = OPCS Classification of Interventions and Procedures code used to identify the coded clinical entry.

| | 2019 | Remain | | |
|---|---|---|---|---|
| | | Yes (+) | No (-) | Total |
| **eR2Ra** | Yes (+) | 214,613 | 21,333 | 235,946 |
| | No (-) | 128,470 | 19,270 | 147,740 |
| | Total | 343,083 | 40,603 | 383,686 |
| | **2020** | **Remain** | | |
| | | Yes (+) | No (-) | Total |
| **eR2Ra** | Yes (+) | 177,852 | 18,283 | 196,135 |
| | No (-) | 97,119 | 17,606 | 114,725 |
| | Total | 274,971 | 35,889 | 310,860 |
| | **2021** | **Remain** | | |
| | | Yes (+) | No (-) | Total |
| **eR2Ra** | Yes (+) | 208,449 | 19,989 | 228,438 |
| | No (-) | 115,111 | 19,075 | 134,186 |
| | Total | 323,560 | 39,064 | 362,624 |

| | 2019 | Remain | | |
|---|---|---|---|---|
| | | Yes (+) | No (-) | Total |
| **eR2Rab** | Yes (+) | 301,018 | 30,176 | 331,194 |
| | No (-) | 42,065 | 10,427 | 52,492 |
| | Total | 343,083 | 40,603 | 383,686 |
| | **2020** | **Remain** | | |
| | | Yes (+) | No (-) | Total |
| **eR2Rab** | Yes (+) | 249,964 | 28,636 | 278,600 |
| | No (-) | 25,007 | 7,253 | 32,260 |
| | Total | 274,971 | 35,889 | 310,860 |
| | **2021** | **Remain** | | |
| | | Yes (+) | No (-) | Total |
| **eR2Rab** | Yes (+) | 291,576 | 30,847 | 322,423 |
| | No (-) | 31,984 | 8,217 | 40,201 |
| | Total | 323,560 | 39,064 | 362,624 |

**Table S3a**. Contingency tables showing the number of patients meeting criteria for eR2Ra and eR2ab and the corresponding number of patients who remain in hospital over the next 24 hours or do not (were discharged), for the in-patient population at 08.00.

| | 2019 | Remain | | |
|---|---|---|---|---|
| | | Yes (+) | No (-) | Total |
| eR2Ra | Yes (+) | 214,005 | 19,919 | 233,924 |
| | No (-) | 129,543 | 18,465 | 148,008 |
| | Total | 343,548 | 38,384 | 381,932 |
| | **2020** | **Remain** | | |
| | | Yes (+) | No (-) | Total |
| eR2Ra | Yes (+) | 178,709 | 17,343 | 196,052 |
| | No (-) | 98,123 | 17,692 | 115,815 |
| | Total | 276,832 | 35,035 | 311,867 |
| | **2021** | **Remain** | | |
| | | Yes (+) | No (-) | Total |
| eR2Ra | Yes (+) | 211,080 | 19,105 | 230,185 |
| | No (-) | 116,893 | 19,616 | 136,509 |
| | Total | 327,973 | 38,721 | 366,694 |

| | 2019 | Remain | | |
|---|---|---|---|---|
| | | Yes (+) | No (-) | Total |
| eR2Rab | Yes (+) | 299,551 | 28,334 | 327,885 |
| | No (-) | 43,997 | 10,050 | 54,047 |
| | Total | 343,548 | 38,384 | 381,932 |
| | **2020** | **Remain** | | |
| | | Yes (+) | No (-) | Total |
| eR2Rab | Yes (+) | 250,507 | 27,672 | 278,179 |
| | No (-) | 26,325 | 7,363 | 33,688 |
| | Total | 276,832 | 35,035 | 311,867 |
| | **2021** | **Remain** | | |
| | | Yes (+) | No (-) | Total |
| eR2Rab | Yes (+) | 294,260 | 30,038 | 324,298 |
| | No (-) | 33,713 | 8,683 | 42,396 |
| | Total | 327,973 | 38,721 | 366,694 |

**Table S3b**. Contingency tables showing the number of patients meeting criteria for eR2Ra and eR2ab and the corresponding number of patients who remain in hospital over the next 24 hours or do not (were discharged), for the in-patient population at 16.00.

| Population at 00:00 | Not meeting eR2Rab criteria and discharged in subsequent 24 hours | Not meeting eR2Rab criteria and not discharged |
|---|---|---|
| n | 22515 | 97326 |
| **Age in years**: median (IQR) | 60(45-74) | 64(49-77) |
| **Sex (n, %)** | . | |
| Female | 10833 (48.1%) | 45345 (46.6%) |
| Male | 11682 (51.9%) | 51981 (53.4%) |
| **Self-reported ethnicity (n, %)** | | |
| White | 15761 (70.0%) | 70194 (72.1%) |
| Mixed/ Multiple | 411 (1.8%) | 1549 (1.6%) |
| Asian/ Asian British | 2952 (13.1%) | 12194 (12.5%) |
| Black/ African/ Caribbean/ Black British | 1274 (5.7%) | 5857 (6.0%) |
| Other ethnic group | 567 (2.5%) | 2285 (2.3%) |
| Not known | 1550 (6.9%) | 5247 (5.4%) |
| **Co-morbidity count (n, %)** | | |
| None | 6544 (29.1%) | 24873 (25.6%) |
| 1-2 | 10321 (45.8%) | 41401 (42.5%) |
| 3 or more | 5650 (25.1%) | 31052 (31.9%) |
| **Morbidities (n, %)** | | |
| Hypertension | 9168 (40.7%) | 43062 (44.2%) |
| Cerebrovascular disease | 1512 (6.7%) | 10128 (10.4%) |
| Atrial fibrillation | 2947 (13.1%) | 17096 (17.6%) |
| Ischaemic heart disease, angina, myocardial infarct | 3810 (16.9%) | 20962 (21.5%) |
| Diabetes (type 1 and 2) | 4809 (21.4%) | 24368 (25.0%) |
| Asthma | 2644 (11.7%) | 9899 (10.2%) |
| COPD | 1594 (7.1%) | 7255 (7.5%) |
| Interstitial Lung Disease | 24 (0.1%) | 129 (0.1%) |
| Chronic Kidney Disease | 3135 (13.9%) | 16633 (17.1%) |
| Any active Malignancy | 3968 (17.6%) | 17572 (18.1%) |
| Dementia (all types) | 535 (2.4%) | 3413 (3.5%) |
| **English Indices of deprivation** | | |
| 1 | 9448 (42.0%) | 38534 (39.6%) |
| 2 | 4638 (20.6%) | 19841 (20.4%) |
| 3 | 3888 (17.3%) | 16630 (17.1%) |
| 4 | 2200 (9.8%) | 9432 (9.7%) |
| 5 | 1644 (7.3%) | 6914 (7.1%) |
| Missing | 697 (3.1%) | 5975 (6.1%) |
| Regained R2R criteria during stay? (n, %) | N/A | 58609 (60.2%) |
| **Reason for regaining R2R criteria?** | | |
| ICU | | 1727 (1.8%) |
| TAP surgery (72h) | | 263 (0.3%) |
| Lower limb surgery (24h) | | 95 (0.1%) |
| Invasive surgery (24h) | N/A | 579 (0.6%) |
| Acute dependency level (48h) | | 7452 (7.7%) |
| NEWS >3 (24h) | | 20605 (21.2%) |
| O2 treatment (24h) | | 5111 (5.3%) |

| | | |
|---|---|---|
| Intravenous fluids or treatments (24 hours, > tds) | | 27069 (27.8%) |
| GCS < or + 12 (24h) | | 183 (0.2%) |
| EOL care (24h) | | 165 (0.2%) |
| Increased dependency (48h) | | 10290 (10.6%) |

**Table S4. Demographics of patients not meeting R2R criteria on presentation to QEHB in the censor period. Legend.** Data is number (percentage) of patients in a bed at 00:00 who either were or were not discharged in the subsequent twenty-four hours after eR2R assessment. Ethnicity was self-reported. Medical conditions were physician confirmed and checked against admission and linked primary care notes. English Indices of deprivation were calculated using postcode.

### Online Supplement Figures

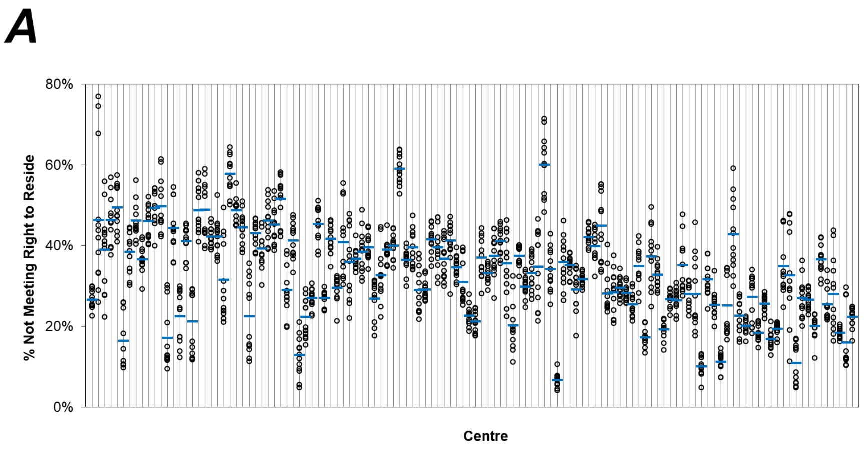


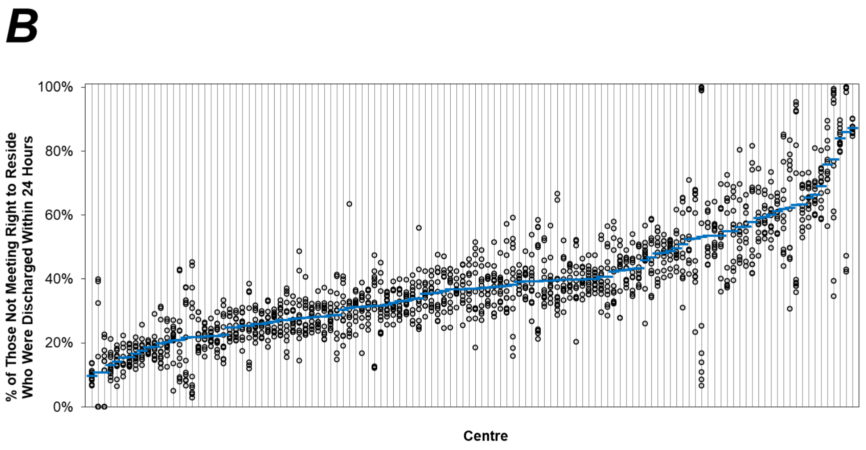


**Figure S1. The proportions of patients with no right to reside (A), and proportions of these that were discharged within 24 hours (B). Analysis by week.**

**Legend:** The proportions of patients not meeting the R2R, and the proportions of these patients that were discharged within 24 hours were extracted from daily reports for each national NHS centre. The weekly mean of each centres values was calculated for each of twelve weeks analysed and plotted as a circle. The mean across the twelve weeks analysed for each centre is plotted as a horizontal line. Centres are arranged in ascending order of the period mean proportion of patients without R2R discharged within 24 hours.

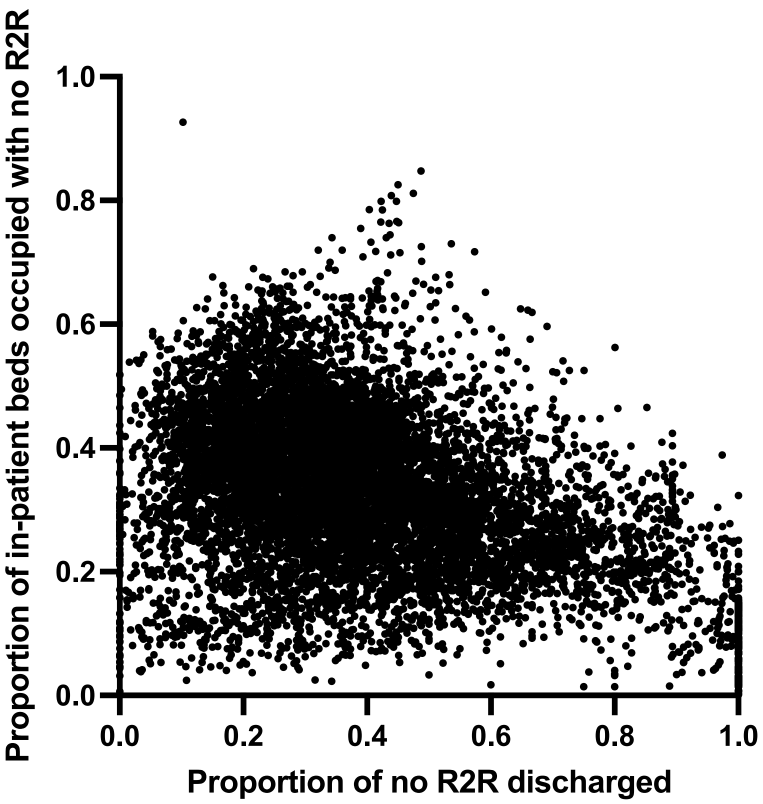


**Figure S2. The proportions of patients with no R2R and of that group the proportion discharged over the next 24 hours**

**Legend:** The proportions of patients not meeting the R2R, and of that group the proportion of patients discharged within 24 hours, reported to SDCS from 29 Nov 2021 – 20 Feb 2022 across 121 centres. Each dot represents result for a single centre-day. The two metrics were associated (slope = -0.21, $p<0.0001$) but the correlation was low ($R^2$=0.12).

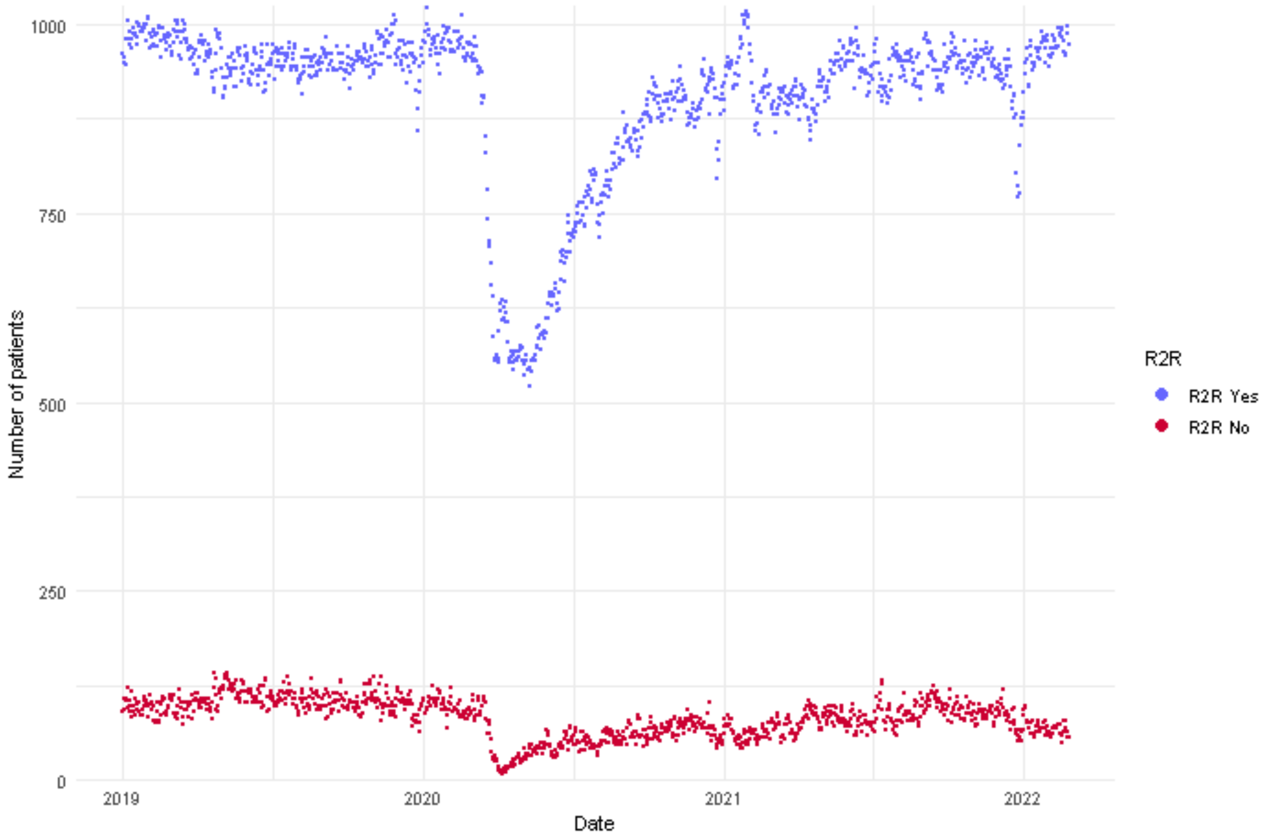


**Figure S3. The number of patients meeting or not meeting eR2Rab criteria 01 Jan 2019 - 31 Dec 2021**

**Legend:** Number of patients with (red dot) or without (blue dot) eR2Rab at 00:00 on each day of 2019-2021. The first COVID-19 admission to QEHB occurred on 1st March 2020. The first wave of the pandemic was associated with significant changes resulting in reduced bed occupancy and the majority of admitted patients had a diagnosis of COVID-19
